# Community health system vital signs and preventable neonatal mortality in Mashonaland West, Zimbabwe: a cluster-randomised controlled trial

**DOI:** 10.64898/2026.08.26.26361392

**Authors:** Meggie Gabida, Eustarckio Kazonga, Kasonde Bowa

## Abstract

Preventable neonatal deaths remain a major public health problem in Zimbabwe, where near-universal antenatal and facility-delivery coverage coexist with a rising neonatal mortality rate. This study evaluated whether institutionalising three core “vital signs” of the community health system (a trained village health worker (VHW) workforce, functional community governance structures, and modified women’s and men’s participatory learning and action groups) reduces preventable neonatal deaths in Mashonaland West Province.

An embedded QUAN (qual) mixed-methods design was used, with a two-arm, parallel-group cluster-randomised controlled trial as the dominant strand. Fifty-two ward-level clusters were randomised 1:1 to the institutionalised community health system package or to standard Ministry of Health and Child Care community services, and 984 pregnant women were enrolled between 1 September 2020 and 31 October 2021, with each mother–infant pair followed to 28 days after delivery, yielding 973 mother–infant pairs for intention-to-treat analysis. The primary outcome was neonatal death within 28 days of life, expressed per 1,000 live births. The primary analysis used a three-level mixed-effects log-binomial regression model with cluster and community-health-worker random intercepts, adjusted for pre-specified covariates. Supervised machine-learning classifiers with leave-one-cluster-out cross-validation, Cox proportional-hazards regression, and multilevel logistic models were fitted as supplementary analyses. An embedded longitudinal process evaluation used key informant interviews and focus group discussions, which were analysed thematically and integrated with the quantitative findings.

The neonatal mortality rate was 44.8 per 1,000 live births in the intervention arm versus 110.1 per 1,000 in the control arm. The adjusted risk ratio for neonatal death was 0.43 (95% CI 0.26–0.70; p < 0.001), a 57% relative reduction, with a number needed to treat of 16 mother–infant pairs (95% CI 11–29). Low birthweight (<2,500 g), birth interval under two years, and low community women’s literacy were the strongest risk factors, while trained VHWs, functional community governance, early antenatal care, and sustained participatory group attendance were independently protective. The women’s and men’s groups were protective in a dose-dependent manner, becoming significant at four or more cycles (about 14 meetings) (adjusted odds ratio 0.71; 95% CI 0.60– 0.85; p = 0.001). A random forest classifier discriminated against neonatal deaths with a cross-validated area under the curve of 0.842 and a sensitivity of 0.912. Qualitative findings converged with the trial results, identifying male engagement, earlier care-seeking, danger-sign literacy, social-network activation, and community death audits as the behavioural and structural mechanisms of change.

Institutionalising the community health system package (trained VHWs, functional governance, early antenatal engagement, and sustained participatory groups) was associated with a substantial reduction in preventable neonatal deaths. The findings suggest that in high-coverage, high-mortality settings, the binding constraint is structural rather than clinical, and that scaling functional community governance and workforce infrastructure in the most disadvantaged communities may accelerate progress toward neonatal survival targets. A one-year follow-up, the rarity of neonatal death, and concurrent national programming that partly reached control clusters are the principal limitations.

**Trial registration:** Pan African Clinical Trials Registry, PACTR202607591142118 (https://pactr.samrc.ac.za/TrialDisplay.aspx?TrialID=PACTR202607591142118); registered retrospectively on 7 July 2026.

## Introduction

The neonatal period remains the most dangerous interval in the human lifespan. Approximately 2.3 million newborns died worldwide in 2024, representing close to half of all under-five deaths, and this burden is concentrated overwhelmingly in sub-Saharan Africa and South Asia [1]. The dominant causes (complications of preterm birth, intrapartum-related events including birth asphyxia, and neonatal infections) are largely amenable to timely and appropriate care, and approximately three-quarters of neonatal deaths occur within the first seven days of life [1,2]. Sustainable Development Goal (SDG) target 3.2 commits all countries to reducing neonatal mortality to 12 or fewer deaths per 1,000 live births by 2030 [3].

Within LMIC health systems, the community tier has remained comparatively underdeveloped relative to facility-based care, despite the fact that the overwhelming majority of maternal, neonatal, and child deaths occur outside tertiary settings [4]. A substantial body of trial evidence indicates that community-based and community-led approaches, notably participatory women’s groups and community health worker programmes, can improve maternal and neonatal outcomes [5,6]. The recurring difficulty has been one of durability rather than efficacy: such approaches are frequently implemented as time-limited projects rather than embedded as financed, supervised, and accountable components of the health system [7].

Zimbabwe exemplifies this pattern. National antenatal care coverage stands at approximately 96% and facility delivery at approximately 86%, yet neonatal mortality has not fallen commensurately, with the national rate estimated at 37 deaths per 1,000 live births in the 2023–24 Demographic and Health Survey, higher than the estimate reported four years earlier [8,9]. Mashonaland West Province, which is predominantly rural, has recorded among the highest provincial rates in the country, reaching 46 per 1,000 live births in 2015 [9]. Village health workers (VHWs) have formed part of Zimbabwe’s primary health care architecture since 1980, following the Alma-Ata Declaration, yet the cadre continues to function on a voluntary basis with modest stipends, uneven geographic distribution, limited supportive supervision, and tenuous connection to community governance bodies [7,10]. The National Community Health Strategy (2020–2025), ratified in 2020, established the policy architecture for recognising the community level as the first tier of the health system [10].

The analytical premise of this study is that neonatal deaths in such settings are patterned by institutional arrangements (how the community tier is financed, staffed, supervised, and held to account) and are therefore amenable to structural intervention rather than reducible to clinical or biological determinants alone [11]. On this premise, the study operationalises three measurable “vital signs” of the community health system, so termed by analogy with clinical vital signs because they indicate whether the system is functioning, accountable, and capable of producing health outcomes: (i) the trained VHW / community health workforce; (ii) functional community governance structures, principally Health Centre Committees (HCCs) and Village Health Committees (VHCs); and (iii) modified women’s and men’s participatory learning and action groups. Each maps onto the World Health Organisation (WHO) health system building blocks and onto the three delays that precede preventable deaths around childbirth [12,13].

No prior study in Zimbabwe has prospectively modelled the effect of simultaneously institutionalising all three community health system vital signs on preventable neonatal deaths; existing work has focused predominantly on single components [14]. This study was designed to fill that gap. The overall aim was to evaluate whether institutionalising the three vital signs, compared with standard community health service delivery, reduces preventable neonatal deaths (attributable principally to preterm birth complications and neonatal infections) among neonates aged 0–28 days in Mashonaland West Province during 2020–2021. The specific objectives were to (1) determine the independent association of each vital sign with neonatal mortality; (2) quantify their individual and combined effects; (3) develop and evaluate a multilevel predictive model of neonatal mortality risk; and (4) identify the strongest predictors and their policy implications. The study tested the null hypothesis (H₀) that institutionalising the community health system vital signs has no effect on reducing preventable neonatal deaths.

## Materials and methods

### Study design

This study used an embedded mixed-methods design, specifically QUAN (qual), in which a two-arm, parallel-group cluster-randomised controlled trial (cRCT) constituted the dominant quantitative strand, and a longitudinal qualitative process evaluation was embedded within it. The design was grounded in a pragmatist paradigm: causal measurement of the intervention’s effect on neonatal death required the internal validity of a controlled trial, while the embedded qualitative strand was needed to explain implementation fidelity, mechanisms of change, and contextual factors such as community perceptions, power dynamics, and structural barriers. The two strands were integrated at the interpretation stage through joint displays and narrative synthesis. The trial was designed as a superiority trial; no equivalence or non-inferiority margin was specified. The study was anchored in four complementary frameworks: the Mosley and Chen child-survival framework [15], the Socio-Ecological Model, the modified Tanahashi health-service-coverage model [16], and Complex Adaptive Systems theory, which together informed variable selection, sampling, data collection, and analysis.

### Study setting

The trial was conducted in Mashonaland West Province, in Zimbabwe’s western region bordering Zambia. The province comprises seven administrative districts (Chegutu, Hurungwe, Kariba, Makonde, Mhondoro-Ngezi, Sanyati, and Zvimba) and contains urban, peri-urban, and rural settlement types, making it broadly representative of the national demographic and geographic profile. According to the 2022 census, the province had a population of 1,893,578, approximately 12.5% of Zimbabwe’s population. It was selected for its high and sustained neonatal event rate (sufficient to power the trial), sparse facility distribution (facilities typically 20–40 km apart, creating natural non-overlapping ward-level clusters and reducing contamination risk), an existing but under-linked VHW workforce, strong provincial and district governance capacity, community engagement designated as a provincial priority, and logistical accessibility from Harare for consistent supervision.

### Participants and eligibility

The unit of randomisation was the community cluster, defined as a ward, which is an established administrative unit corresponding to the catchment area of a single primary health care facility (clinic), each serving approximately 8,000–10,000 people within a radius of 5–8 km and served by a dedicated, non-overlapping network of VHWs. All wards within the province were eligible. The primary target population comprised women of childbearing age (15–49 years) resident in eligible wards, identified during pregnancy and followed to 28 days after delivery. Wards containing or served by a referral hospital were excluded to minimise the confounding effect of tertiary-level care, as were non-permanent residents. Recruitment was conducted at the ward level. VHWs identified pregnant women confirmed at the local clinic, who were then approached at the household level and enrolled following written informed consent.

Clusters were recruited, and pregnant women were identified and enrolled between 1 September 2020 and 31 October 2021; each enrolled mother–infant pair was followed until 28 days after delivery. The trial is registered with the Pan African Clinical Trials Registry (PACTR202607591142118; https://pactr.samrc.ac.za/TrialDisplay.aspx?TrialID=PACTR202607591142118). Registration was completed retrospectively on 7 July 2026, after data collection had ended. The trial was conducted in 2020–2021 as unfunded doctoral research under ethics approval obtained before enrolment (University of Lusaka Institutional Review Board, IORG0010092-082; Medical Research Council of Zimbabwe, MRCZ/A/2558); it was not registered before enrolment because prospective registration was not, at that time, a requirement of the approving ethics committees or the doctoral programme, and the study was subsequently entered under the registry’s initiative to capture trials already in progress or completed. No outcomes, endpoints, eligibility criteria, or analyses were altered after data collection. The authors confirm that all ongoing and related trials for this intervention are registered.

### Randomisation, allocation concealment, and blinding

Of 211 wards assessed for eligibility, 159 were excluded (16 within 5 km of district or mission hospitals providing secondary care, and 143 peripheral wards designated as buffer zones to prevent contamination). The remaining 52 clusters were randomised 1:1 to the intervention or control arm (26 per arm). An independent biostatistician, not involved in recruitment or data collection, generated the allocation sequence using restricted (blocked) randomisation with fixed blocks of four clusters (two per arm), stratified by district, in Stata version 14. Block sizes were concealed, and the sequence was stored in a password-protected file and released to the field team only once recruitment at a site was ready to begin, ensuring allocation concealment. Blinding of participants and providers was not possible because the intervention involved visible cluster-level changes; however, data collectors recording trial outcomes were blinded to allocation, all data were coded and anonymised before analysis, and analysts remained blinded to group allocation. No breaches of blinding were reported.

Contamination was minimised through geographic separation of wards (20–40 km of sparsely populated terrain), strict VHW boundary fidelity monitored through supervision logs, and the 143 buffer-zone wards. Spillover was measured directly: at each follow-up visit, women in control clusters were asked a standardised screening question about exposure to intervention content, and the proportion reporting any exposure was used as a secondary process outcome.

### Intervention and comparator

Control clusters received the standard package of community health services in accordance with national Ministry of Health and Child Care (MOHCC) policy, comprising routine VHW home visits, standard antenatal and postnatal referral pathways, and existing community governance structures operating without the structured institutionalisation package. Intervention clusters received the same routine services plus the institutionalised package of three vital signs. First, the VHW / community health workforce component: one VHW per village (or per approximately 100 households) received 92 hours of training in essential newborn care, danger-sign recognition, VHW–facility linkage, and postnatal home visitation, and established networks of neighbouring households (“community health forums”) that themselves received 24 hours of training on the Community Essential Health Package.

Second, the modified women’s and men’s participatory learning and action (WLA/MLA) component adapted the women’s group model previously implemented in Bolivia, Nepal, and Malawi [17–19] to the Zimbabwean context and, recognising the patriarchal setting, deliberately included men. VHWs guided groups through five community-mobilisation cycles over the trial period, spanning problem identification and prioritisation, locally led planning, community-wide implementation, evaluation and resilience-building, and a final social-accountability cycle incorporating community-based maternal, neonatal, and perinatal death audits. To complement the group cycles, VHWs conducted scheduled home visits during pregnancy and in the early postnatal period, promoting the WHO-recommended eight antenatal care contacts, skilled birth attendance, clean cord care, thermal care and kangaroo mother care, early and exclusive breastfeeding, and danger-sign recognition and referral. Third, the community governance component revitalised HCCs and VHCs and defined clear linkages with the formal health system, with over 4,000 VHWs, HCC and VHC members, health facility teams, and district health teams oriented to data-driven decision-making, community monitoring, and the use of community scorecards.

Intervention fidelity was assessed prospectively across the five domains of the NIH Behaviour Change Consortium Treatment Fidelity Framework [20] (design, provider training, delivery, receipt, and enactment). A composite cluster-level fidelity score (Fi) was computed as the weighted mean of standardised domain scores; clusters with Fi < 0.70 were flagged for remediation, and a pre-specified per-protocol sensitivity analysis restricted the intervention arm to high-fidelity clusters. A theme-based drama (“*Mwana Wedu*”) planned as a mobilisation tool was suspended because of COVID-19 restrictions and is not reported here.

### Outcomes

The primary outcome was neonatal death within 28 days of life, expressed as a rate per 1,000 live births and analysed by intention to treat. The trial endpoint was all-cause neonatal mortality; preventability was invoked conceptually rather than as an outcome criterion, on the basis that the causes principally targeted by the intervention (preterm birth complications and neonatal sepsis) are preventable through timely community and facility-level care. Deaths were ascertained through household surveillance and confirmed by verbal autopsy or facility records, and sub-classified as early (0–6 days) or late (7–27 days) neonatal deaths. Verbal autopsy data were coded to WHO and ICD-10 standards; at least two physicians independently assigned causes of death, with inter-rater reliability assessed using Cohen’s kappa and discrepancies adjudicated by a third senior physician. Pre-specified secondary outcomes included timely care-seeking for danger signs, immediate newborn-care practices, maternal service uptake (≥4 and ≥8 antenatal contacts, postnatal care), functionality of community governance structures, neonatal morbidity, the contamination rate in control clusters, and the composite fidelity score.

### Sample size

The sample size was calculated for the primary outcome using the method of Hayes and Bennett for cluster-randomised trials, assuming a baseline neonatal mortality rate, a two-sided 5% significance level with 80% power, a between-cluster coefficient of variation (k) of 0.25, and a target relative reduction consistent with prior participatory-group trials in Nepal and Bolivia. Allowing for 10% loss to follow-up, this yielded a minimum of 489 mother–infant pairs per arm (978 in total) across 52 clusters (26 per arm). The empirical intracluster correlation coefficient observed in the trial (approximately 0.026 at the cluster level) was consistent with the assumptions underlying this calculation. For the qualitative strand, sampling was guided by thematic saturation, with at least two focus group discussions per major subgroup per arm and 5–8 key informant interviews per stakeholder category, continued until no new themes emerged.

### Statistical analysis

Quantitative data were analysed using SPSS version 29 and Stata version 14. The primary analysis employed a three-level mixed-effects log-binomial regression model to estimate the risk ratio (RR) for neonatal death, accounting for the hierarchical structure of individuals nested within community health workers (CHWs) and CHWs nested within clusters, with random intercepts specified at both levels. Intervention effects are presented as adjusted risk ratios (aRRs) with 95% confidence intervals (CIs).

Covariate selection followed a pre-specified causal framework informed by a directed acyclic graph (DAG). Prespecified confounders (district, baseline neonatal mortality, and Helping Babies Breathe/Helping Babies Survive training) were included in all models regardless of statistical significance. Prespecified mediators (antenatal care visits and facility delivery) and colliders (postnatal complications) were excluded a priori to avoid biased estimates. Maternal age and parity were retained as precision variables where they improved model precision. A secondary univariable screening step (p < 0.05) was used solely to identify additional candidate covariates not specified in the DAG, thereby limiting data-driven model selection. To account for multiple testing across the 46 candidate predictors, p-values were adjusted using the Benjamini–Hochberg false discovery rate procedure.

Missing data were handled using complete-case analysis when missingness was less than 5%. When missingness exceeded 5%, multiple imputation by chained equations was performed under the missing-at-random assumption, with sensitivity analyses comparing estimates from the imputed and complete-case datasets.

Three prespecified supplementary analyses were conducted. First, multilevel logistic regression was used to compare weighted two- and three-level random-intercept models incorporating village health worker (VHW) variables and interaction terms. Model fit was evaluated using the Akaike information criterion (AIC) and Bayesian information criterion (BIC).

Second, an exploratory machine-learning analysis evaluated four supervised classifiers (logistic regression, random forest [21], decision tree, and support vector machine) implemented in scikit-learn version 1.0.2 in Python 3.8 using the full analytic sample (973 participants, including 68 neonatal deaths [6.99%]). To preserve the clustered data structure and prevent information leakage, model performance was assessed using leave-one-cluster-out (LOCO) cross-validation [22], with all preprocessing conducted within each training fold. Class weighting addressed outcome imbalance, and hyperparameters were optimised using nested cross-validation. Predictive performance was evaluated using the area under the receiver operating characteristic curve (AUC), sensitivity, specificity, positive and negative predictive values, the Brier score, and Hosmer– Lemeshow calibration. Variable importance was assessed using mean decrease in impurity, permutation importance, and SHAP (Shapley additive explanations) values. These analyses were exploratory, internally validated, and were not used to estimate the causal effect of the intervention.

Third, a Cox proportional hazards model was fitted to identify community health system factors associated with the hazard of neonatal death during the 28-day follow-up. The proportional hazards assumption was assessed using Schoenfeld residuals.

Qualitative interviews were transcribed verbatim, translated where necessary, and analysed thematically in NVivo using an iteratively developed coding framework, cross-theme matrices, and narrative synthesis. Descriptive summaries of qualitative findings were generated in Stata version 14.

### COVID-19 protocol adaptations

The following deviations from the approved trial protocol occurred and are reported here in full. The recruitment and follow-up timeline shifted by approximately one year relative to the approved protocol (Version 2, 28 January 2020), which had planned enrolment over September 2019 to September 2020; actual recruitment ran from 1 September 2020 to 31 October 2021 owing to COVID-19–related delays to fieldwork, while the trial design, arms, eligibility criteria, outcomes, and analyses were otherwise unchanged. The COVID-19 pandemic necessitated protocol adaptations during 2020–2021, including removal of the day-3 postnatal home visit in line with national guidelines, suspension of the group drama and large in-person gatherings, and increased reliance on telephone follow-up. These adaptations may have led to under-ascertainment of early neonatal outcomes and introduced recall bias and are recognised as limitations; however, reduced inter-cluster mobility during the pandemic also plausibly improved fidelity and reduced contamination. Additional adaptation-specific fidelity and contamination indicators were collected, and all deviations were documented in the trial registry and CONSORT flow.

Reporting followed the CONSORT 2010 statement [23] and its extension for cluster-randomised trials [24] (S1 Checklist).

### Ethics statement

The study received ethical approval from the University of Lusaka Institutional Review Board (Ref. IORG0010092-082) and the Medical Research Council of Zimbabwe (MRCZ/A/2558). Permission was obtained from the MOHCC provincial and district health offices. Written informed consent was obtained from all participating mothers, including consent for their minor infants; consent forms were provided in English and Shona. Participation was voluntary, without monetary compensation, and participants could withdraw at any time. Confidentiality and anonymity were maintained using study-unique identifiers, and data were stored securely for the period required by the University of Lusaka and MRCZ ethics requirements. All COVID-19 public health precautions were observed throughout data collection.

## Results

### Trial profile and baseline characteristics

The trial ran from 1 September 2020 to 31 October 2021, with each enrolled mother–infant pair followed to 28 days after delivery. Of 211 wards assessed, 52 were randomised (26 per arm). No cluster failed to receive its allocation, and none was lost to follow-up. A total of 984 pregnant women were enrolled (intervention 483; control 501); 11 (1.1%) could not participate because they required their husbands’ consent (2 in the intervention arm and 9 in the control arm), leaving 973 mother–infant pairs for intention-to-treat analysis (intervention 481; control 492), representing 99.6% follow-up in the intervention arm and 98.2% in the control arm. Among the 973 pregnancies analysed, there were 91 stillbirths (intervention 35; control 56), yielding 882 live births (intervention 446; control 436) and 68 neonatal deaths, of which 47 (69.1%) were early, and 21 (30.9%) were late neonatal deaths (Fig 1).

**Fig 1.**
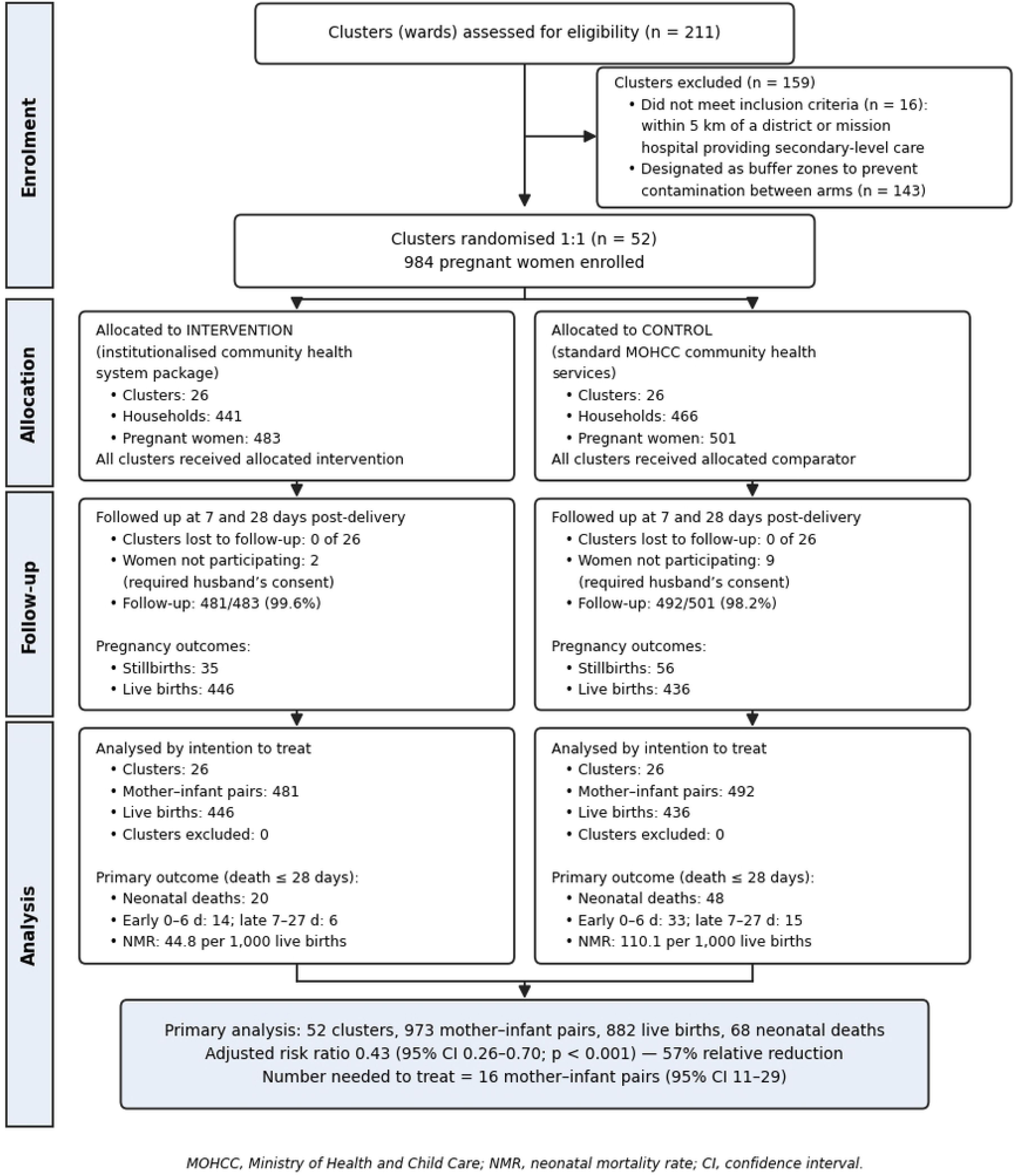
CONSORT flow of clusters and participants through the trial. Of 211 wards assessed for eligibility, 159 were excluded (16 within 5 km of a hospital providing secondary-level care; 143 designated as buffer zones to prevent contamination between arms), leaving 52 clusters randomised 1:1. No cluster failed to receive its allocation or was lost to follow-up. All 973 mother–infant pairs were analysed intention-to-treat within their originally assigned clusters.

Baseline characteristics were well balanced between arms, with all standardised mean differences below 0.10 (maximum 0.048; Table 1). Households and mothers were broadly comparable across arms in residence, wealth quintile, maternal age, marital status, religion, education, employment, parity, birth interval, and polygamy (all p > 0.05). One clinically relevant imbalance favoured the intervention arm: low birthweight (<2,500 g) was less common in the intervention arm (12.9%) than in the control arm (18.3%; p = 0.020).

**Table 1:** Baseline characteristics by trial arm.

| Characteristic | Intervention | Control | SMD |
| --- | --- | --- | --- |
| Maternal age, years (mean (SD)) | 26.4 (6.4) | 26.7 (6.4) | 0.048 |
| Primiparous, % | 28.3 | 29.1 | 0.018 |
| Secondary education or higher, % | 42.7 | 41.9 | 0.016 |
| Married, % | 91.2 | 90.8 | 0.014 |
| ≥4 ANC visits, % | 66.8 | 67.3 | 0.011 |
| Multiple pregnancy, % | 1.8 | 2.1 | 0.023 |
| Previous neonatal death, % | 8.7 | 8.2 | 0.019 |
| Facility delivery, % | 78.4 | 77.9 | 0.012 |
| Skilled birth attendant, % | 76.2 | 75.8 | 0.010 |
| Caesarean section, % | 8.7 | 9.1 | 0.014 |
| Wealth quintile 4–5, % | 35.2 | 34.8 | 0.008 |
| Clean water access, % | 62.4 | 61.9 | 0.010 |
| Improved sanitation, % | 48.7 | 49.3 | 0.010 |

ANC, antenatal care; SD, standard deviation; SMD, standardised mean difference. All $SMD < 0.10$ indicates good balance.

### Primary outcome: neonatal mortality at 28 days

In the intervention arm, 20 neonatal deaths occurred among 446 live births (neonatal mortality rate 44.8 per 1,000 live births), compared with 48 deaths among 436 live births in the control arm (110.1 per 1,000 live births). The crude rate ratio was 0.41. After adjustment for the pre-specified covariates (birthweight, birth interval, maternal age, community governance functionality, and socioeconomic status), the adjusted risk ratio for neonatal death in the intervention arm relative to control was 0.43 (95% CI 0.26–0.70; p < 0.001), corresponding to a 57% relative reduction in neonatal mortality. The number needed to treat to prevent one neonatal death was 16 mother– infant pairs (95% CI 11–29). The stability of the estimate before and after adjustment is consistent with the baseline balance achieved through randomisation. Stillbirth rates were also lower in the intervention arm (72.8 versus 113.8 per 1,000 total births).

### Community, service-delivery, and governance characteristics

Intervention and control arms differed markedly on community and service-delivery characteristics reflecting the intervention (Table 2). Fully functional community governance structures were reported by 82.7% of women in intervention communities versus 34.3% in control communities (p < 0.0001). Participation in WLA/MLA group meetings reached 97.1% in intervention areas versus 12.6% in control areas (p < 0.0001). VHW home visits (72.6% versus 39.2%), VHWs trained in newborn care (67.4% versus 39.2%), early antenatal booking before 12 weeks (60.9% versus 42.7%), ≥8 antenatal contacts (54.3% versus 42.3%), household danger-sign recognition (69.4% versus 50.8%), and kangaroo mother care at home (72.1% versus 41.1%) were all significantly higher in intervention communities (all p ≤ 0.05). By contrast, national-programme indicators such as skilled birth attendance did not differ between arms (76.5% versus 77.9%; p = 0.62), confirming clear differentiation between national and intervention-specific activities.

**Table 2:** Selected community and service-delivery characteristics by trial arm (n = 973).

| Characteristic (proportion reporting) | Intervention (n=481) | Control (n=492) | *p*-value |
| --- | --- | --- | --- |
| Fully functional community governance | 82.7% | 34.3% | <0.0001 |
| WLA/MLA group participation | 97.1% | 12.6% | <0.0001 |
| Formal CHW contact | 96.0% | 73.4% | <0.0001 |
| VHW trained in newborn care | 67.4% | 39.2% | <0.0001 |
| VHW home visit | 72.6% | 39.2% | <0.0001 |
| Social networks present | 74.0% | 36.0% | <0.0001 |

| Characteristic (proportion reporting) | Intervention (n=481) | Control (n=492) | p-value |
| --- | --- | --- | --- |
| Early ANC booking (<12 weeks) | 60.9% | 42.7% | <0.0001 |
| ≥8 ANC contacts | 54.3% | 42.3% | <0.0001 |
| Household danger-sign recognition | 69.4% | 50.8% | <0.0001 |
| Kangaroo mother care at home | 72.1% | 41.1% | 0.045 |
| Skilled birth attendant | 76.5% | 77.9% | 0.619 |

ANC, antenatal care; CHW, community health worker; VHW, village health worker; WLA/MLA, women's and men's learning and action.

### Risk and protective factors

Of the 46 candidate predictors examined, 22 remained significantly associated with neonatal death after Benjamini–Hochberg correction. In bivariate analysis, low birthweight (<2,500 g) was the strongest single correlate of neonatal mortality (r = 0.72), followed by birth complications (r = 0.37) and non-functional community governance (r = 0.24). In the multivariable logistic regression (Table 3), low birthweight (adjusted odds ratio [aOR] 5.10; 95% CI 3.98–6.54), birth interval under two years (aOR 3.85; 95% CI 3.22–4.60), low community women’s literacy (aOR 4.05; 95% CI 3.18–5.16), birth complications (aOR 2.75; 95% CI 2.29–3.31), low socioeconomic status (aOR 2.55; 95% CI 1.96–3.32), and non-functional community governance (aOR 1.80; 95% CI 1.50– 2.16) were independently associated with increased odds of neonatal death (all p < 0.0001). Protective factors included early antenatal care before 12 weeks (aOR 0.36; 95% CI 0.21–0.59), household danger-sign recognition (aOR 0.35; 95% CI 0.22–0.56), female infant sex (aOR 0.52; 95% CI 0.36–0.76), and the presence of social networks (aOR 0.43; 95% CI 0.28–0.66) (all p < 0.0001).

**Table 3.** Multivariable logistic regression: factors independently associated with neonatal mortality, Mashonaland West Province, 2020–2021.

| Variable | Adjusted OR (95% CI) | *p*-value |
| --- | --- | --- |
| Low birthweight <2,500 g | 5.10 (3.98–6.54) | <0.0001 |
| Birth interval <2 years | 3.85 (3.22–4.60) | <0.0001 |
| Low community women's literacy | 4.05 (3.18–5.16) | <0.0001 |
| Birth complications | 2.75 (2.29–3.31) | <0.0001 |
| Low socioeconomic status | 2.55 (1.96–3.32) | <0.0001 |
| Polygamous marriage | 1.88 (1.46–2.43) | <0.0001 |
| Non-functional community governance | 1.80 (1.50–2.16) | <0.0001 |
| Rural residence | 1.15 (1.03–1.29) | 0.008 |
| Female infant sex | 0.52 (0.36–0.76) | <0.0001 |
| Early ANC before 12 weeks | 0.36 (0.21–0.59) | <0.0001 |
| Household danger-sign recognition | 0.35 (0.22–0.56) | <0.0001 |
| Social networks present | 0.43 (0.28–0.66) | <0.0001 |
| WLA/MLA participation ($\geq 4$ cycles) | 0.71 (0.60–0.85) | 0.001 |
| WLA/MLA participation (<3 cycles) | 4.80 (1.95–11.80) | <0.0001 |

ANC, antenatal care; OR, odds ratio; CI, confidence interval; WLA/MLA, women's and men's learning and action. Adjusted for pre-specified directed acyclic graph-informed covariates.

Participation in four or more WLA/MLA cycles was associated with significantly lower odds of neonatal death (aOR 0.71; 95% CI 0.60–0.85; p = 0.001), a 29% reduction, whereas completing fewer than three cycles was associated with a nearly fivefold increase in risk (aOR 4.80; 95% CI 1.95–11.80; p < 0.0001), indicating a dose-response relationship consistent with a cumulative participatory social-change mechanism.

### Component effects and synergy (secondary estimand)

In adjusted log-binomial models estimating component-specific risk ratios, trained VHWs (aRR 0.271; 95% CI 0.16–0.38), fully functional community governance (aRR 0.327; 95% CI 0.21–0.45), and social networks (aRR 0.336; 95% CI 0.23–0.44) were each strongly protective, corresponding to relative risk reductions of 72.9%, 67.3%, and 66.4% respectively. Early antenatal care (aRR 0.366; 95% CI 0.27–0.46) and household danger-sign recognition (aRR 0.424; 95% CI 0.32–0.53) were similarly protective, whereas skilled birth attendance alone was not significantly associated with reduced neonatal death (p = 0.37). The combined effect of components was estimated formally using interaction terms within the three-level model rather than by multiplying component risk ratios. Two interactions were statistically significant: VHW presence with a short birth interval (aOR 0.42; 95% CI 0.24–0.73; p = 0.002) and VHW newborn-care training with early antenatal care (aOR 0.38; 95% CI 0.21–0.69; p = 0.001), indicating that the joint presence of these components was associated with a greater reduction in the odds of neonatal death than either component alone. Communities with fully institutionalised community governance achieved approximately 43% greater reduction in neonatal deaths than those with only partial implementation.

### Multilevel and survival models

Among the multilevel models, the weighted three-level random-intercept model incorporating VHW variables and interactions (Model 4) provided the best fit, with the lowest AIC (764.35) and BIC (866.88) of all models tested, compared with the null model (AIC 1362.0) and the two-level covariate model (AIC 810.06). In this model, short birth interval (aOR 2.94; 95% CI 1.28–6.78) and low community women’s literacy (aOR 2.35; 95% CI 1.25–4.41) increased the odds of neonatal death, while VHW presence (aOR 0.64; 95% CI 0.44–0.94), VHW newborn-care training (aOR 0.51; 95% CI 0.33–0.77), and VHW home visits (aOR 0.68; 95% CI 0.48–0.97) were protective. Household-level clustering explained 28.6% of the variance in the three-level model.

The Cox proportional-hazards model (68 events; omnibus χ² = 65.594, df = 5, p < 0.0001) identified fully functional community governance (hazard ratio [HR] 0.073; 95% CI 0.037–0.141; p < 0.001) and women’s and men’s group participation (HR 0.097; 95% CI 0.053–0.180; p < 0.001) as the most strongly protective community health system vital signs, followed by VHW training in newborn care (HR 0.503; p < 0.05) and enhanced social networks among pregnant mothers (HR 0.426; p < 0.05). Non-functional governance showed no significant association (HR 1.097; p > 0.05).

### Exploratory predictive modelling

Under leave-one-cluster-out cross-validation, the random forest classifier achieved the strongest discrimination (AUC 0.842; 95% CI 0.816–0.860), with sensitivity 0.912, specificity 0.803, and the lowest Brier score (0.0369); it correctly identified 62 of the 68 neonatal deaths (Table 4). The decision tree performed comparably (AUC 0.830) and offers a transparent, deployable rule structure, while the support vector machine (AUC 0.686) and logistic regression (AUC 0.615) underperformed, consistent with the non-linear, interaction-dependent structure of the data. Positive predictive values were modest across all models, a structural consequence of the 6.99% outcome prevalence, and calibration, as assessed by the Hosmer–Lemeshow test, was adequate for the logistic regression and support vector machine models but not for the two tree-based models. Feature-importance analyses ranked short birth interval, low birth weight, family influence, infant sex, and household danger-sign recognition as among the strongest predictors. These analyses are exploratory, internally validated, and do not estimate the causal effect of the intervention; external validation is required before any deployment as a clinical decision-support tool.

**Table 4.** Leave-one-cluster-out cross-validated performance of four predictive classifiers (n = 973; 68 neonatal deaths).

| Model | AUC | Sensitivity | Specificity | Brier |
| --- | --- | --- | --- | --- |
| Random forest | 0.842 | 0.912 | 0.803 | 0.0369 |
| Decision tree | 0.830 | 0.912 | 0.790 | 0.0387 |
| Support vector machine | 0.686 | 0.750 | 0.670 | 0.0398 |
| Logistic regression | 0.615 | 0.611 | 0.622 | 0.0423 |

*AUC, area under the receiver operating characteristic curve. Metrics reflect aggregated out-of-fold predictions; class weighting applied to all algorithms.*

### Intervention fidelity

Across all 26 intervention clusters, the mean composite fidelity score was 0.964 (SD 0.09; range 0.84–0.97), and no cluster fell below the pre-specified remediation threshold. Fidelity was temporally stable over the 12-month trial period, with no significant trend (β = 0.002; p = 0.78), attributed to refresher training at the start of each learning cycle. There was perfect containment of WLA/MLA group meetings (0% in control clusters), although the composite spillover indicator in control clusters was 45.6%, driven mainly by VHW newborn-care training (37.9%) reaching control areas through concurrent national programming and informal information-sharing. This co-intervention is addressed as a limitation below.

### Qualitative findings and mixed-methods integration

Key informant interviews and focus group discussions across all seven districts (138 participants) yielded four themes: understanding of institutionalisation; availability coverage of community health system components; accessibility of community health services; acceptability, utilisation, and effective coverage. Institutionalisation was frequently equated with government ownership and VHW geographic coverage, with broader systemic components (governance, social accountability, and community participation) underemphasised, particularly at district level. Participants described the 2020 National Community Health Strategy as a transformative catalyst for investment in previously deprioritised components, but reported uneven governance revitalisation, with HCCs supported more consistently than ward-level VHCs. Community leaders described acute geographic barriers, with some communities more than 30 km from the nearest clinic, and persistent exclusion of artisanal miners and households in remote areas.

The modified women’s and men’s groups emerged as the central mechanism for demand creation, with participants emphasising that the inclusion of men shifted household decision-making in a patriarchal setting. A recurrent account linked prenatal naming of the infant to emotional bonding, earlier antenatal booking, and care-seeking readiness; correspondingly, 70% of neonatal deaths in intervention areas occurred in infants who had not been named, and absence of a name was independently associated with mortality (aOR 2.40; 95% CI 1.52–3.78; p < 0.0001). A gendered feeding norm was also documented, in which male infants were given complementary foods before the recommended 6 months. Community leaders described a transition in governance structures toward active accountability, including community-based death audits and emergency transport mobilisation. Across all four themes, the qualitative and quantitative strands converged: the behavioural and structural mechanisms articulated by informants (male engagement, earlier care-seeking, danger-sign literacy, social-network activation, and community accountability) corresponded to the quantitatively estimated protective effects of the intervention components, and community structures were credited with maintaining service delivery through the pandemic.

## Discussion

In this cluster-randomised controlled trial in a high-burden rural province of Zimbabwe, institutionalising three community health system vital signs (a trained VHW workforce, functional community governance, and modified participatory women’s and men’s groups) was associated with a 57% relative reduction in neonatal mortality (aRR 0.43; 95% CI 0.26–0.70), with one additional neonatal death prevented for roughly every 16 mother–infant pairs in communities receiving the full package. The finding was consistent across the primary log-binomial analysis, the multilevel and survival models, the exploratory machine-learning strand, and the embedded qualitative evaluation, and each of the three components carried an independent protective association while producing larger, super-additive effects in combination.

These results bear on a discrepancy that has characterised Zimbabwe’s maternal and newborn health profile: high levels of service contact have not translated into commensurate newborn survival [9,25]. Where antenatal and delivery coverage are already high, the residual mortality burden is unlikely to be resolved by further increments in coverage alone. The absence of a significant association between skilled birth attendance and neonatal survival in this trial is consistent with that reading, although it must be interpreted cautiously: this study did not measure the content or quality of intrapartum care, and the null result should not be construed as evidence that skilled attendance is unimportant. Rather, the pattern suggests that once contact is established, survival depends on whether the surrounding system can act on what that contact reveals (through referral, audit, transport, and feedback between clinic and community). This maps onto the three-delays framework: the VHW component operates on the decision to seek care and the journey to care, whereas governance operates on the adequacy of care received [13].

The trial extends the evidence base on participatory women’s groups in two respects. Previous cluster-randomised trials evaluated such groups largely in isolation, without an accompanying governance component [5,6,17,18]. Here, the groups were protective in a dose-dependent fashion, reaching significance only among participants completing four or more cycles (about 14 meetings) (aOR 0.71) and attenuating below that threshold, a gradient consistent with a mechanism of gradual normative and behavioural change requiring sustained exposure, rather than an immediate effect of allocation. Second, the magnitude of the combined effect is compatible with, though it does not prove, the hypothesis that governance structures convert community mobilisation into an adaptive feedback loop; cross-trial comparisons remain observational, and differences in effect size may equally reflect population heterogeneity, design variation, or differing follow-up duration. Supporting this interpretation, functional community governance emerged as the strongest community-level structural predictor of survival in the Cox model (HR 0.073), and community structural variables retained predictive value of a magnitude comparable to established biological predictors in both the multilevel and machine-learning analyses - a signal that facility-based models, which do not observe these variables, cannot detect.

The distributional findings carry policy weight. Mortality risk clustered in the poorest, most geographically isolated, and least literate communities, which were also those with the weakest governance functionality and sparsest VHW coverage. Interpreted structurally, this co-location is not incidental: it reflects the accumulated geography of underinvestment in the community tier, and it argues for allocating resources according to structural deficit rather than distributing them uniformly. The practical consequences are the formal recognition and remuneration of VHWs within the primary health care workforce rather than their continued treatment as volunteers [26]; the devolution of genuine operational and budgetary authority to VHCs, with a defined reporting line to district health teams; multi-year rather than project-cycle funding for participatory groups, given the dose-response gradient observed here; and the incorporation of governance-functionality indicators into district performance monitoring alongside the coverage indicators that currently dominate it [7,10]. Priority should fall on the settings least served to date, including artisanal mining settlements and households situated more than 30 km from the nearest facility.

### Strengths and limitations

Strengths include the prospective cluster-randomised design with concealed, stratified, blocked allocation and near-complete follow-up; a pre-specified, causally informed analytic structure with correction for multiple testing; triangulation across quantitative, machine-learning, survival, and qualitative strands; and prospective, multidomain fidelity monitoring. Several limitations temper the findings. First, the follow-up was 1 year, so the possibility that the dose-dependent participatory-group effect would strengthen as normative change matured could not be assessed. Second, neonatal death is a rare event (6.99% prevalence), which limits the precision of interaction estimates and raises the risk of overfitting in the exploratory models; the machine-learning analyses are internally cross-validated only and require external validation before any deployment. Third, and most importantly for effect estimation, Zimbabwe’s phased national rollout of a revised community health strategy during 2020–2021 reached some control clusters (composite spillover 45.6%, driven by VHW training), a co-intervention that would tend to attenuate rather than inflate the observed effect, making the estimate a conservative lower bound. Fourth, COVID-19 adaptations, including the removal of the day-3 visit and the suspension of the group drama, may have led to under-ascertainment of early outcomes and to compressed participation in some clusters. Fifth, generalisability is constrained: the trial was conducted in one province, and replication in other provinces and in comparable LMIC settings [27] is required before the findings can be considered transferable. Recruitment through facility-confirmed pregnancy may also have underrepresented the most geographically isolated households, which would bias the distributional analysis toward the null and render the equity conclusions conservative rather than overstated. Cause-of-death classification relied partly on facility-register abstraction, where verbal autopsies could not be completed, introducing scope for misclassification. Finally, this study identifies low birthweight and low women’s literacy as strong predictors but did not directly evaluate interventions targeting them; such interventions require intersectoral action beyond the community health system.

## Conclusions

Institutionalising the three community health system vital signs (a trained VHW workforce, functional community governance, and sustained participatory women’s and men’s groups, together with the early antenatal engagement they generate) was associated with a substantial reduction in preventable neonatal mortality in Mashonaland West Province. The components were not interchangeable: partial implementation produced markedly smaller gains, and the significant workforce-by-governance interaction indicates that their value is realised jointly. Where service coverage is already high, these results suggest that the remaining constraint on newborn survival lies less in extending contact than in constructing the governance and workforce infrastructure through which contact becomes effective care. This has direct operational implications for Zimbabwe’s National Community Health Strategy, which already provides the policy architecture: the evidence presented here identifies which components to prioritise within it and where to concentrate them. Four research priorities follow: a three-to-five-year follow-up to establish whether the effect is sustained as governance matures and as participatory groups accumulate exposure; a formal economic evaluation of cost per death averted; external validation of the predictive model in at least two comparable settings before any individual-level deployment; and implementation research on how institutionalisation can be achieved at scale under resource constraints.

## Data Availability

Data cannot be shared publicly because of ethical and legal restrictions. The consent obtained from participants, and the conditions attached to the ethical approvals granted for this trial (University of Lusaka Institutional Review Board, IORG0010092-082 Medical Research Council of Zimbabwe (MRCZ/A/2558) restrict the use of individual participant-level records to the study investigators. The dataset comprises household-level maternal and neonatal records, including neonatal deaths, drawn from small, geographically defined rural wards, and carries a residual risk of re-identification that de-identification alone does not eliminate. All aggregate data underlying the results reported in the manuscript are contained within the manuscript. Requests for access to de-identified data may be sent to the Medical Research Council of Zimbabwe or to the corresponding author and will be considered subject to ethics committee approval.

## Acknowledgments

The authors thank the women of Mashonaland West Province who participated in the study; the Provincial Nursing Officer and district health teams for facilitating implementation; the village health workers and community governance structures; the data collection and supervision teams; and the postgraduate coordination office at the University of Lusaka. The authors are grateful to all community members, leaders, and health facility teams who supported the trial under challenging conditions, including those imposed by the COVID-19 pandemic.

## Supporting information

**S1 Checklist - CONSORT 2010 checklist for cluster-randomised trials:** Completed checklist of standard CONSORT items together with the extension items for cluster-randomised trials, indicating where each item is reported in the manuscript.

